# Potential applications of the rapid COVID-19 antibody test kit screening in comparison to the RT-PCR in patients and personnel at the Department of Obstetrics and Gynecology

**DOI:** 10.1101/2021.07.16.21259725

**Authors:** Amarin Narkwichean, Wittaya Jomoui, Rarinthip Boonpradit, Wipada Laosooksathit, Tanawin Nopsopon, Krit Pongpirul

**Author notes:** Corresponding Author: Krit Pongpirul, M.D., M.P.H., Ph.D. Associate Professor, Department of Preventive and Social Medicine, Faculty of Medicine, Chulalongkorn University, Thailand; Department of International Health, Johns Hopkins Bloomberg School of Public Health, Baltimore, MD, USA. Synopsis: The article describes the application of the rapid COVID-19 antibody test kit to screen both patients (admitted for operation/procedure) and medical personnel in O&G department.

## Abstract

**Objective:** To explore potential applications of the rapid antibody test for COVID-19 screening, in comparison to RT-PCR, for emergency obstetric and gynecological procedures, and medical personnel in the Department of Obstetrics and Gynecology.

**Methods:** A cross-sectional study was conducted in expected 290 participants: 230 patients and 60 medical staff, during the four-month national COVID-19 outbreak period (Aug – Sep 2020, and Dec 2020 - Jan 2021). All participants underwent both rapid antibody tests and RT-PCR (at admission for patients).

**Results:** A total of 270 participants completed the study. Fever and URI symptoms were present in 6/210 patients (2.8%) while one patient (0.5%) had a history of traveling to a high-risk area. However, only two (1%) asymptomatic patients had positive IgM results. Concerning the medical personnel, 10% fell into the ‘patient under investigation (PUI)’ category. 4/60 (6.7%) IgM positive was observed in the staff cohort in which 3/4 came from non-PUI participants. Neither participant had RT-PCR positive demonstrating a 1.9% total false positive rate.

**Conclusion:** Rapid point-of-care antibody test can be used to screen either a pregnant coming for delivery, a patient who requires urgent/emergency operative procedures, or medical personnel, at least in the defined lower-prevalence COVID-19 situation.

**Trial registration:** This study was approved by the Institutional Review Board of Srinakharinwirot University (IRB SWUEC119/2563F) and was registered at the Thai Clinical Trials Registry (TCTR20210613001).

## Introduction

Since the first report from Wuhan China in December 2019 [1], the novel coronavirus (SARS-CoV-2) pandemic has become a global catastrophe. The SARS-CoV-2 infection causes severe acute respiratory syndrome yet a majority (>80%) of symptomatic infections have the mild disease [2] whereas approximately one-third of the infections are asymptomatic [3]. It is expected that more than half of all transmissions came from asymptomatic transmission either pre-symptomatic or asymptomatic individuals [4]. The gold standard of investigation is the real-time RT-PCR to detect the viral RNA from the nasopharyngeal swab. A systematic review observed a pooled sensitivity at 87.8% (95% CI, 81.5%, and 92.2%) for an initial RT-PCR test [5]. False-negative real-time PCR testing is notable especially in asymptomatic patients, which can be reduced by performing multiple tests [6]. Other limitations include high costs, taking time to get the result, and requirements of experienced staff and sophisticated equipment. Moreover, real-time PCR is not available in every institution, thus may lengthen the result turn-around time.

Thailand has experienced few episodes of national outbreaks, the latest one recently from March 2021. During each epidemic, all hospitals operate following governmental policy. A majority of outpatient and elective operation services are suspended to reduce the crowdedness of the area. Hospital visitors and staff are screened utilizing temperature scan and their clinical history. A ‘patient under investigation (PUI)’, or a person at risk of having the virus, is isolated to undertake the RT-PCR investigation. The utmost priority is to prevent intra-hospital clusters as health facilities i.e., negative pressure rooms, staff, personal protective equipment (PPE) are limited. Nonetheless, emergency and urgency procedures are to be maintained with the highest precautions. The obstetrics and gynecology services anticipate various urgent/emergent conditions. Obstetric procedural practices, both complicated and uncomplicated deliveries as well as interventions for miscarriage, must be retained whereas patients with either a gynecologic malignancy or pelvic mass complication unavoidably require urgent/immediate surgical intervention. The local recommendation states that in the epidemic area, all cases coming for surgical intervention do require pre-admission RT-PCR test, otherwise in lower prevalence areas, the PUI criteria are used for the RT-PCR investigation. Concerns are raised regarding the reliability of the PUI criteria especially in cases of asymptomatic women. In the obstetrics and gynecology atmosphere, most patients and health personnel are young and healthy, thus are more likely to be asymptomatic or have mild symptoms. Moreover, although RT-PCR is available, it is rather challenging to obtain the results in all cases particularly those patients who are either multiparous or in an advanced stage of labor. As such, PPE and other equipment would be quickly consumed to prevent intrahospital clusters.

A rapid point-of-care antibody test could be a useful screening tool before performing an obstetric and gynecological procedure. Baiya’s rapid COVID-19 IgG/IgM test kit (Baiya Phytopharm, Bangkok), developed in early 2020 by the faculty of Pharmacology, Chulalongkorn University, is a rapid point-of-care antibody test in either serum or plasma, utilizing ‘the lateral flow immunoassay’ technique. The result is read within 15 minutes. Patient’s antibodies, both IgG and IgM, to SARS-CoV-2 spike protein are detected when they were bound to the kit’s conjugate membrane coated with recombinant antigen protein produced from the plant in the laboratory. The lateral flow immunoassay has been proved to be effective in literature with 88.66% sensitivity and 90.63% specificity [7]. The kit post-manufacturer evaluation in the laboratory utilizing 51 samples of confirmed positive case and 151 samples control observed 94% (48/51) sensitivity and 98% (147/150) specificity of either positive IgM or IgG result. Due to a significant proportion of asymptomatic infection mentioned above, particularly a higher rate of asymptomatic infection (49-68%) in pregnant women reported in the literature [8]. The study was conducted i) to evaluate diagnostic values of the Baiya’s rapid IgG/IgM test kit as a point-of-care COVID-19 screening in comparison to the RT-PCR in patients and medical personnel, and ii) to evaluate the seroprevalence of the COVID-19 in.

## Materials and Methods

This cross-sectional study was performed at the Department of Obstetrics and Gynecology, Srinakharinwirot university hospital, during the period between August – September 2020 and December 2020 - January 2021. A total of 290 participants were expected; i) 230 patients and pregnant women visiting the hospital for either an emergency medical procedure or an elective unpostponable surgery including vaginal delivery, cesarean section, uterine curettage for miscarriage, and exploratory laparotomy, ii) 60 medical personnel, comprising physicians, nursing staffs, technicians, in the department. Participants were excluded when they denied or could not complete both the rapid test and RT-PCR testing. The study received ethical approval from the institutional review board (SWUEC119/2563F). Informed consent was obtained from the participants. The study received funding from the Health System Research Institute (HSRI), Thailand (HRSI63-132) and was non-commercially supported for the Baiya’s rapid covid-19 IgM/IgG test kits by the Faculty of Pharmacology, Chulalongkorn University.

Demographic data and risks associated with COVID-19 infection including i) history of closed contact with confirmed COVID-19 patient, ii) history of traveling to a high-risk area, iii) clinical symptoms consisting of fever, cough, rhinorrhea, sore throat, dyspnea, or anosmia were recorded by a physician. A blood sample was taken either by fingertip collection or venipuncture. It was dropped into a test kit for 1-2 drops followed by 2-3 drops of dilution buffer solution and waited for 15 minutes to analyze the test result. The rapid test result was read by a trained physician. Subsequently, the nasopharyngeal swab was obtained. The RT-PCR for SARS-CoV-2 was performed on the ABI7500 real-time PCR machine (ThermoFisher, Waltham, USA) using a commercial kit (Sansure Biotech Inc, China). This assay detects two specific SARS-CoV-2 genes including ORF-1ab and N genes (lower limit of detection; LOD = 0.2 copies/uL). To ensure appropriate specimen collection, the RNase P gene was used as an internal control (IC). The cut-off cycle threshold (ct) was assigned at less than 40 for interpretation with ‘detectable’. The RT-PCR tests were analyzed by a molecular pathologist (WJ) who was blinded from the rapid test results.

During the epidemic, the physician and health personnel received PPE for treating PUI/confirmed COVID-19 patients appropriate to the national center for disease controls standard, i.e. cover-all suit, apron, cap, face-shield, N95-respirator, gloves, and leg covers for PUI patients performing in the negative pressure environment. If the patient had no PUI risk, the health personnel used a surgical mask, gloves, and face-shield with surgical gown when performing a surgical procedure as suggested by the hospital infectious control unit. The airborne-protection-grade PPE (research-funded) was also provided to cover blood tests and swab collection procedures in all participants. In the emergency event or patients needed an emergency medical procedure, the nasopharyngeal swab was done after the procedure (the doctors contacted with unverified patients were wearing PPE during the medical procedure).

### Statistical analysis

The sample size, of patients visiting the hospital for medical procedures, was calculated using a statistic formula and a prevalence of COVID-19 infection in pregnancy previously reported by Sutton D. et al (2020) [9]. A total participant (plus 20%) of 230 patients were expected. Other 60 participants who are health personnel were anticipated. Data are presented using descriptive statistics. We had planned to evaluate the sensitivity, specificity, and predictive values of the Baiya’s rapid covid-19 IgM/IgG test kit, in comparison to the real-time RT-PCR, if the seroprevalence was allowed.

## Results

During the 4-month study period, a total of 232 pregnant and patients admitted to the hospital for an operative procedure participated in the study. Twenty-two participants were excluded, all due to the real-time PCR was not performed, either by participants withdrawing consent or technical/clinical issues. The median age of the remaining 210 participants was 29 years (IQR 25-36 years old). The majority were Thai (82.9%) while the others were Burmese (18/210, 8.6%), Cambodian (13/210, 6.2%), and Laotian (5/210, 2.4%). More than 95% of Burmese and Cambodians were undocumented immigrant workers (30/31). More than 80% of participants were admitted to the hospital for childbirth delivery either vaginal delivery (57.6%) or cesarean section (25.7%). The cesarean section cases included both elective (admitted 12-24 hours prior) or emergency operation. There were 14% of patients admitted for gynecological surgery including oncological operations and emergency laparotomy/laparoscopic surgeries (ruptured ectopic pregnancy, torsion ovarian cyst) (Table 1).

**Table 1:** The demographic data of patients and pregnant women.

| **Demography; N=210** | **Value** |
| --- | --- |
| **Age (years): Median [IQR]** | **29 [25, 36]** |
| **Reason for admission: N (%)** |  |
| Vaginal delivery | 121 (57.6%) |
| Cesarean section | 54 (25.7%) |
| Gynecologic oncology/Gynecology major operation | 29 (13.8%) |
| Curettage for obstetric/gynecologic conditions | 6 (2.9%) |
| **Race: N (%)** |  |
| Thai | 174 (82.9%) |
| Burmese | 18 (8.6%) |
| Cambodian | 13 (6.2%) |
| Laotian | 5 (2.4%) |
| **History of travel to high-risk area (without mask); N (%)** | **1 (0.5%)** |
| **History of contact with patient under investigation / confirmed positive; N (%)** | **0 (0.0%)** |
| **Health care worker; N (%)** | **2 (1.0%)** |
| **Presence of symptoms\*; N (%)** | **6 (2.8%)** |
| Fever 37.5 C | 6 (2.8%) |
| Cough | 2 (1.0%) |
| Rhinorrhea | 1 (0.5%) |
| Sore throat | 1 (0.5%) |

\*A patient could have multiple symptoms

There were six symptomatic participants (3.2%). Fever, cough, rhinorrhea, and sore throat were observed in six (3.2%), two (1%), one (0.5%), and one (0.5%) participant(s), respectively. Neither had contact with a confirmed COVID-19 case. Only one patient (0.5%) reported that she had been visiting a declared high-risk area. Overall, there were two participants (1%) with the positive rapid antibody test result, both having only IgM positive. These two patients neither had URI symptoms, had been traveling to the high-risk area, nor had contact with a confirmed COVID-10 patient. The real-time PCR for SARS-CoV-2 was negative in both patients.

The first case was a pregnant woman, aged 36-40, who was admitted due to labor pain with two-centimeter cervical dilatation. Her fetus had been estimated large since during her antenatal care. Her obstetrician had initially planned for labor augmentation. Nonetheless, upon arrival, her rapid antibody test was positive. She was moved into the negative-pressure labor suite while waiting for the urgent RT-PCR result (expected three-hour waiting time). Her obstetrician then decided to perform a cesarean section upon obtaining the RT-PCR result to limit contamination. Both mother and fetus were well following the operation and received routine postpartum care.

The second case was a woman, aged 61-65, diagnosed with early-stage endometrial cancer admitted for the surgical staging operation. Her physician decided to perform the procedure as it was believed that the surgery could provide the patient with the best disease prognosis. Nonetheless, her operation was postponed for one week because of the positive IgM rapid test result on admission. Repeat real-time PCR on the second admission before the operation was confirmed negative, thus, the procedure and care were performed in the standard-setting (Table 2).

**Table 2:** Characteristics of participants with positive rapid IgM/IgG antibody test.

| Case | Status | Indication | Age /Sex | Risk factors |  |  | Rapid Ab test |  | RT-PCR | Intervention |
| --- | --- | --- | --- | --- | --- | --- | --- | --- | --- | --- |
|  |  |  |  | Travel to a high-risk area | Hx of contact | Symptoms | IgM | IgG |  |  |
| 1 | Patient | Surgical staging | 61-65 (F) | No | No | None | (+) | (-) | Negative | Operation postponed for 1 week<br>Repeated RT-PCR - negative prior operation |
| 2 | Patient | Cesarean section (Emergency) | 36-40 (F) | No | No | None | (+) | (-) | Negative | Requested for emergency RT-PCR<br>Operation delayed for 3 hours |
| 3 | HCW | Volunteer to research | 31-35 (F) | No | No | None | (+) | (-) | Negative | Leave and self-quarantine until RT-PCR result known (1 Day) |
| 4 | HCW | Volunteer to research | 36-40 (M) | No | No | None | (+) | (-) | Negative | Leave and self-quarantine until RT-PCR result known (1 Day) |
| 5 | HCW | Volunteer to research | 25-30 (F) | No | No | Rhinorrhea<br>No fever | (+) | (-) | Negative | Leave and self-quarantine for 2 days<br>Until symptoms deceased /<br>RT-PCR result known (1 day)<br>then self-monitoring for 14 days |

A total of 60 medical personnel participated in the study from December 2020 to January 2021, which was the nationwide second outbreak of COVID-19 in Thailand. The median age was 28 years old (Interquartile range 24.8-38). The majority were women (78.3%) and nursing staff (60%). Just above a quarter were physicians including interns, residents, clinical fellows, and consultants. The cohort also included 5 pre-licensed final-year medical students who had just been relocated from a provincial hospital which was considered a high-risk area. Also, there was one resident who just visited the declared high-risk area and one intern who had contacted a confirmed COVID-19 patient while working in the other hospital after hours. Six medical personnel had upper respiratory tract symptoms. Overall, 12 (20%) medical staff were classified as patients under investigation for COVID-19 including one physician who had contacted with confirmed COVID-19 case, five medical students who traveled from the high-risk area, and six medical personnel who had upper respiratory tract symptoms (Table 3). The rapid antibody test results were positive IgM-only antibodies in four participants (6.8%). None of the medical personnel tested had a positive real-time PCR for SARS-CoV-2 (Table 2).

**Table 3:** The demographic data of medical personnel.

| **Demography; N=60** | **Value** |
| --- | --- |
| **Age (years): Median [IQR]** | 28 [24.8, 38] |
| **Female N (%)** | 47 (78.3%) |
| **Role N (%)** |  |
| Physician | 17 (28.4%) |
| Registered / Practical Nurse | 36 (60.0%) |
| Final-year medical students | 5 (8.3%) |
| Technician/ Laboratory scientist | 2 (3.3%) |
| **Recent traveled to or worked in high-risk area; N (%)** | **6 (10%)** |
| Number of IgM/IgG rapid test positive | 0 (0%) |
| **History of direct/indirect contact with patient under investigation /confirmed positive; N (%)** | **1 (1.7%)** |
| Number of IgM/IgG rapid test positive | 0 (0%) |
| **Presence of symptoms (fever, cough, rhinorrhea, sore throat); N (%)** | **6 (10%)** |
| Number of IgM/IgG rapid test positive | 1/6 (16.7%) |

## Discussion

The major challenge during the pandemic is to balance between prevention of intrahospital SARS-CoV-2 transmission and provision of patient-centered care for emergency/urgency conditions. Standard/complicated obstetric and indicated gynecologic operation services must be maintained. Pregnant women are more likely to either requiring invasive ventilation support or be admitted to ICU when compared to reproductive-age non-pregnant women. Obstetric complications such as premature labor and maternal death are increased in those having the COVID-19 infection. However, data demonstrates that they are less likely to show symptoms such as fever, dyspnea, and myalgia [10], thus they are potentially a silence spreader when visiting the hospital. Our major research questions were to evaluate the potential benefit of using the rapid point-of-care antibody test for screening SARS-CoV-2 in comparison with the real-time PCR as well as to investigate the actual seroprevalence of patients in the obstetrics and gynecology department.

To date, there was no definite practice guideline for screening obstetrics and gynecology patients before admitted to the hospital [11]. The Royal Thai College of Obstetricians and Gynecologists (RTCOG) recommended that the RT-PCR should be performed on admission in every case in the high-risk area setting. Concerns have been raised regarding the universal RT-PCR screening because of its high cost, time to get results, and availability in every hospital. An initial RT-PCR false-negative result, particularly in an asymptomatic patient, has been reported in the literature [6, 12]. Zullo and colleagues suggested screening SARS-CoV-2 antibodies utilizing a rapid antibody test in pregnant women before receiving care in both inpatient and outpatient settings. The positive either IgM or IgG antibody result is subsequently confirmed by performing a nasopharyngeal swab for real-time PCR [13]. Nonetheless, more studies are needed to support the use of rapid antibody tests, especially when there were differences in terms of countries’ prevalence and test-kit products. We selected Baiya’s rapid COVID-19 IgG/IgM test kit, developed by Phoolcharoen and colleagues, which had been tested for efficiency both in the laboratory and in few other clinical trials during this study period.

Our research shows that though there were two nationwide outbreak episodes, the seroprevalence of patients with positive IgM/IgG antibodies was only 1% (2/210). Neither of these two was symptomatic, thus did not meet the PUI criteria, and were not eligible for RT-PCR according to the Thai government policy. On the contrary, six pregnant participants were having fever (and URI symptoms) demonstrating a 3.4% (6/175) rate of intrapartum fever. Intrapartum fever is not uncommon in obstetric practice, reported between 3-7% in the literature [14]. This condition generated great concern about whether patients were having COVID-19 infection. In the pandemic, the RT-PCR is suggested with limitations regarding access and duration of the result as previously described. The rapid test could guide the management concerning the prioritization of PPE equipment and facilities, i.e., negative pressure theatre or cohort ward, which shall be limited during the pandemic. However, the rapid antibody test should be interpreted with caution also due to the odds of false-negative results especially during the early clinical course [5, 13]. In our current practice, we perform both rapid antibody test and RT-PCR in symptomatic patients (fever, URI symptoms) at admission for a patient who requires an urgent/emergency procedure including childbirth delivery.

The research also studied the seroprevalence of SARS-Cov-2 antibodies among the medical personnel, who were considered healthy and active, thus possess a risk to become a ‘silence spreader’. We included 74.07% of all staff in the department. All frontline people including residents, interns, nursing staff, and technicians participated in the study. While only 10% of consultants took part in the study. The Thai government also considers healthcare workers with URI symptoms as one of the PUI criteria. In the current study cohort, 21.6% (12/60) were at risk for COVID-19 infection. About half of these high risk were symptomatic (6/20) while the others were either indirectly contacting the COVID-19 patient or recently traveling to the high-risk area. Contracting the disease from outside the organization either by working- or non-working related is one of the greatest threats of the department in which, despite precaution, the incidences still occurred [15]. Nonetheless, we observed 6.7% (4/60) positive IgM from the rapid test in the study cohort while the majority of the positive (3/4) came from participants without PUI risks. Ultimately, although all positive IgM participants were real-time PCR negative, the authors believe that using a rapid antibody test could improve the detection rate of healthcare workers who are suspected of infection adding up from using the PUI criteria alone.

RT-PCR is the current standard of COVID-19 detection. In our study, the results from the rapid antibody test have corresponded with the RT-PCR with a 1.9% false-positive rate (5/270; range 1% to 6.8% in patients and medical personnel, respectively). During the study period, there was no standard immunoassay for COVID-19 available in our hospital, thus it is unable to confirm the serologic status of the positive rapid test kit. An epidemiological study concerning seroprevalence of COVID-19 antibodies done by Nopsopon T et al. (2020) utilizing Baiya’s rapid COVID-19 IgG/IgM kit in medical personnel and patients undergoing an operation in 52 community hospitals across the country observed 4.5% overall positive antibody results (12.1% and 3.7% in patients and medical personnel, respectively) [16]. While another study from the same group observed 0.8% IgM positive (7/844) seroprevalence in medical personnel in Ranong, a COVID-19-free province (low-risk area). All seven medical personnel were confirmed real-time PCR negative [17]. In both studies, the authors found that there was no correlation between positive rapid antibody test results with PUI risks and symptoms. Moreover, the difference in timing of positive test results between the antigen (RT-PCR) and antibody (point-of-care rapid antibodies kit or standard immunoassay) tests should be considered. The onset and duration of the detection are distinct, for example, RT-PCR will detect with the highest chance in week 1 following the symptom onset while antibodies (mainly IgM) are commonly found in the 2nd week [12, 13]. In asymptomatic participants, it is rather difficult to pinpoint the onset of the disease, thus may pass the detection period of real-time PCR. Nonetheless, further study is needed to confirm the serologic status of the positive rapid antibodies test before concluding that the result is not a false positive one.

Concerning the participants with positive IgM antibody results (Table 2), the oncologic gynecological operation was rescheduled for one week following two consecutive negative RT-PCR results, the second test was just one day before her surgery. The three healthcare workers took their leave and quarantine while waiting for the RT-PCR result. The doctor who was having URI symptoms at the time of the tests returned to work few days after once all symptoms ceased. Lastly, the pregnant woman who had come for delivery, the urgent RT-PCR was then requested in which, three hours later, the result was known to be undetectable. Her labor had been expected to be obstructed since her admission thus she needed to wait for few hours before her cesarean section. We anticipated that all five participants were anxious and stressed while waiting for their RT-PCR result. The situation could have been worse if the hospital did not have an in-house PCR facility which could make up to three days to get the result. Although the real-time PCR was negative in the pregnant woman, her doctor decided to use full PPE for the cesarean section. Nonetheless, while a positive rapid test result caused concern and anxiety to both personnel and patient, a negative result reassured them significantly. More studies are needed to clinically obtain sensitivity and false-negative rates to ensure the safety of using the rapid antibody test to screen both personnel and patients, especially in the pregnant population. In the meantime, PPE should be offered for the emergency surgical procedure during the pandemic. Though the rapid test can guide and facilitate the management of PPE and the negative room more efficiently.

In conclusion, the rapid point-of-care antibody test can be used as either a screening tool for patients who require urgent/emergency operative procedures or a seroprevalence survey on medical personnel on service. In the defined low-prevalence sub-population i.e., pregnant or gynecologic patients, the test kit was relatively accurate in terms of the negative result as compared with RT-PCR with a false positive rate of 1%. While the seroprevalence of medical personnel demonstrates 6.7% positive IgM results who required further investigation (real-time PCR) and management (self-quarantine). The rapid antibody test ensures, while reducing anxiety, patients and healthcare workers at the point of care and facilitate the management of PPE more efficiently, at least in the low to moderate COVID-19 prevalence area. Nevertheless, more data is needed to evaluate the sensitivity and accuracy of the test kit especially in the upscaled pandemic and in the vaccine era in comparison with standard RT-PCR and serologic immunoassays, respectively.

## Supporting information

Study Protocol (English)

STROBE Checklist

## Data Availability

All data relevant to the study are included in the article. De-identified data of this study could be shared upon request.

## Author Contributions

**Conceptualization:** Amarin Narkwichean, Krit Pongpirul

**Data curation:** Amarin Narkwichean, Wittaya Jomoui, Rarinthip Boonpradit, Wipada Laosooksathit

**Formal analysis:** Amarin Narkwichean, Rarinthip Boonpradit, Krit Pongpirul

**Investigation:** Amarin Narkwichean, Tanawin Nopsopon, Krit Pongpirul

**Methodology:** Tanawin Nopsopon, Krit Pongpirul

**Project administration:** Krit Pongpirul

**Resources:** Krit Pongpirul

**Software:** Tanawin Nopsopon, Krit Pongpirul

**Supervision:** Krit Pongpirul

**Validation:** Amarin Narkwichean, Tanawin Nopsopon, Krit Pongpirul

**Visualization:** Amarin Narkwichean

**Writing – original draft:** Amarin Narkwichean, Krit Pongpirul

**Writing – review & editing:** Amarin Narkwichean, Tanawin Nopsopon, Krit Pongpirul, Wittaya Jomoui, Rarinthip Boonpradit, Wipada Laosooksathit

## Acknowledgments

The study received full financial support from the Health System Research Institute (Thai government agencies), Thailand. The authors also thank the Baiya Phytopharm company (Bangkok, Thailand) for providing the Baiya’s Rapid IgG/IgM antibodies test kits to be used in the project.

## Reference

[1] Sun P, Lu X, Xu C, Sun W, Pan B: Understanding of COVID-19 based on current evidence. J Med Virol 2020;92(6): 548–551.

[2] Wu Z, McGoogan JM: Characteristics of and Important Lessons From the Coronavirus Disease 2019 (COVID-19) Outbreak in China: Summary of a Report of 72 314 Cases From the Chinese Center for Disease Control and Prevention. Jama 2020;323(13): 1239–1242.

[3] Oran DP, Topol EJ: The Proportion of SARS-CoV-2 Infections That Are Asymptomatic : A Systematic Review. Ann Intern Med 2021;174(5): 655–662.

[4] Johansson MA, Quandelacy TM, Kada S, Prasad PV, Steele M, Brooks JT, et al.: SARS-CoV-2 Transmission From People Without COVID-19 Symptoms. JAMA Netw Open 2021;4(1): e2035057.

[5] Jarrom D, Elston L, Washington J, Prettyjohns M, Cann K, Myles S, et al.: Effectiveness of tests to detect the presence of SARS-CoV-2 virus, and antibodies to SARS-CoV-2, to inform COVID-19 diagnosis: a rapid systematic review. BMJ Evidence-Based Medicine 2020: bmjebm-2020-111511.

[6] Arevalo-Rodriguez I, Buitrago-Garcia D, Simancas-Racines D, Zambrano-Achig P, Del Campo R, Ciapponi A, et al.: False-negative results of initial RT-PCR assays for COVID-19: A systematic review. PLOS ONE 2020;15(12): e0242958.

[7] Li Z, Yi Y, Luo X, Xiong N, Liu Y, Li S, et al.: Development and clinical application of a rapid IgM-IgG combined antibody test for SARS-CoV-2 infection diagnosis. Journal of Medical Virology 2020;92(9): 1518–1524.

[8] Yanes-Lane M, Winters N, Fregonese F, Bastos M, Perlman-Arrow S, Campbell JR, et al.: Proportion of asymptomatic infection among COVID-19 positive persons and their transmission potential: A systematic review and meta-analysis. PLoS One 2020;15(11): e0241536.

[9] Sutton D, Fuchs K, D’Alton M, Goffman D: Universal Screening for SARS-CoV-2 in Women Admitted for Delivery. N Engl J Med 2020;382(22): 2163–2164.

[10] Allotey J, Stallings E, Bonet M, Yap M, Chatterjee S, Kew T, et al.: Clinical manifestations, risk factors, and maternal and perinatal outcomes of coronavirus disease 2019 in pregnancy: living systematic review and meta-analysis. BMJ 2020;370: m3320.

[11] Yeo KT, Oei JL, De Luca D, Schmölzer GM, Guaran R, Palasanthiran P, et al.: Review of guidelines and recommendations from 17 countries highlights the challenges that clinicians face caring for neonates born to mothers with COVID-19. Acta Paediatrica 2020;109(11): 2192–2207.

[12] Sethuraman N, Jeremiah SS, Ryo A: Interpreting Diagnostic Tests for SARS-CoV-2. JAMA 2020;323(22): 2249–2251.

[13] Zullo F, Di Mascio D, Saccone G: Coronavirus disease 2019 antibody testing in pregnancy. American Journal of Obstetrics & Gynecology MFM 2020;2(3, Supplement): 100142.

[14] Towers CV, Yates A, Zite N, Smith C, Chernicky L, Howard B: Incidence of fever in labor and risk of neonatal sepsis. Am J Obstet Gynecol 2017;216(6): 596.e591-596.e595.

[15] Sikkema RS, Pas SD, Nieuwenhuijse DF, O’Toole Á, Verweij J, van der Linden A, et al.: COVID-19 in health-care workers in three hospitals in the south of the Netherlands: a cross-sectional study. The Lancet Infectious Diseases 2020;20(11): 1273–1280.

[16] Nopsopon T, Pongpirul K, Chotirosniramit K, Hiransuthikul N: COVID-19 Antibody in Thai Community Hospitals. medRxiv 2020: 2020.2006.2024.20139188.

[17] Nopsopon T, Pongpirul K, Chotirosniramit K, Jakaew W, Kaewwijit C, Kanchana S, et al.: Seroprevalence of Hospital Staff in Province with Zero COVID-19 Cases. medRxiv 2020: 2020.2007.2013.20151944.

