## Supplementary material for "Potential applications of the rapid COVID-19 antibody test kit screening in comparison to the RT-PCR in patients and personnel at the Department of Obstetrics and Gynecology": Study Protocol (English)

1. **Scientific Title:**

Sensitivity, specificity, and accuracy of the Baiya’s COVID-19 Rapid Test in comparison to Real-time PCR for diagnosis of the infection in emergency obstetrics and gynecology patients and medical personnel

2. **Public Title:**

COVID-19 antibody testing utilizing the Rapid IgM/IgG antibodies test kit in comparison to the RT-PCR results in patients, admitted to hospital for obstetrics/operative procedure, and personnel at the department of Obstetrics and Gynaecology

3. **Investigators**

Principle Investigator	Dr Amarin Narkwichean, M.D., Ph.D.

Affiliation	Clinical Lecturer, Department of Obstetrics and Gynecology

Faculty of Medicine, Srinakharinwirot University, Thailand

Co-Investigator

SWU – Site

1. Dr Wipada Laosooksathit, M.D.

Affiliation	Assistant Professor, Department of Obstetrics and Gynecology

Faculty of Medicine, Srinakharinwirot University, Thailand

2. Dr Wittaya Jomoui, Ph.D.

Affiliation	Assistant Professor, Department of Pathology

Faculty of Medicine, Srinakharinwirot University, Thailand

3. Dr Rarinthip Boonpradit, M.D.

Affiliation	Department of Obstetrics and Gynecology

Faculty of Medicine, Srinakharinwirot University, Thailand

CU-Site (Baiya’s COVID-19 Rapid Antibodies Test kit)

1. Prof Dr Narin Hiransuthikul, M.D., Ph.D.

Affiliation	Administrative Board, Chulalongkorn University, Thailand

**(Vice President for Strategic Monitoring and Assessment, Planning, Budgeting and Well-being)

2. Dr Krit Pongpirul, M.D., M.PH., Ph.D.

Affiliation	Associate Professor,

Department of Preventive and Social Medicine

Faculty of Medicine, Chulalongkorn University, Thailand

3. Dr Waranyoo Phoolcharoen, Ph.D.

Affiliation	Associate Professor, Faculty of Pharmaceutical Sciences

Chulalongkorn University, Thailand

4. Dr Suthira Taychakhoonavudh, Ph.D.

Affiliation	Assistant Professor, Faculty of Pharmaceutical Sciences

Chulalongkorn University, Thailand

5. Dr Tanawin Nopsopon, M.D.

Affiliation	Lecturer, Department of Preventive and Social Medicine

Faculty of Medicine, Chulalongkorn University, Thailand

4. **Research Funding**

Health System Research Institute (HSRI) funding scheme 2020

HSRI 63-132 *(623700 THB or approx. 17800 USD)*

5. **Sponsor**

The Strategic Wisdom and Research Institute, Srinakharinwirot University

20^th^ Floor, Prof. Dr. Saroch Buasri’s building, Srinakharinwirot Prasarnmitr Campus

114, Sukhumwit 23, Wattana, Bangkok, 10110, Thailand.

6. **Sponsor (IRB) ID**

SWU-EC 119/2563F (Approval date 5^th^ May 2020)

7. **Conflict of Interest**

PI declares that there is no conflict of interest on the following

- Financial conflicts of interest (study sponsorship, funds/grants or any financial support
- Using a company’s resources for personal gain
- Working for a competing business
- Holding shares in a. company which might be influenced by the publication

8. **Primary study site**

Single center – Department of Obstetrics and Gynecology, HRH Maha Chakri Sirindhorn Medical Center (MSMC), Srinakharinwirot University Hospital

9. **Protocol Synopsis**

A descriptive study conducted to evaluate sero-prevalence of SAR-CoV-2 antibodies in comparison to the RT-PCR testings in i) 230 pregnant women and patients visiting the hospital for an emergency/urgency obstetrics and operative procedure, and ii) 60 medical personnel at the department of Obstetrics and Gynaecology, Srinakharinwirot University Hospital.

10. **Study Objectives**

Primary objective

1. To investigate diagnostic values (i.e., sensitivity, specificity, and accuracy) of the Baiya’s Rapid Covid-19 IgM/IgG test kit to detect COVID-19 infection in comparison to the standard RT-PCR in ...
   i. patients and pregnant women visiting the hospital for either an emergency medical procedure or an elective unpostponable surgery including vaginal delivery, cesarean section, uterine curettage for miscarriage, and exploratory laparotomy
   ii. medical personnel having contact with patient at the department of obstetrics and gynecology

Secondary objective

1. To evaluate seroprevalence of COVID-19 in both patients (as above) and personnel at the department of obstetrics and gynecology during the pandemic.
2. To compare the efficacy/efficiency of the Baiya’s Rapid Covid-19 IgM/IgG test kit and the national ‘patient under investigation’ criteria (mainly patient’s history) to screen COVID-19 infection in the study population

11. **Study design**

Clinical research

Diagnostic test (Cross-Sectional Studies)

Single population study

12. **Subject selection and allocation**

Inclusion criteria

1. Patients or pregnant women admitted to the hospital for an operative procedure
   including vaginal delivery, cesarean section delivery, uterine curettage for obstetric complications, and emergency exploratory laparotomy, during the COVID-19 outbreak
2. medical personnel, who are having contacts with patients, of the obstetrics and gynecology department

Exclusion criteria

1. A participant who denies or cannot complete both the rapid antibody test or RT-PCR

13. **Sample size calculation**

For patient participants – referred to Sutton D, et al. (2020) concerning the incidence of positive COVID-19 cases in pregnant women coming for delivery at 14.4% (p=0.144)

By using the formula as following
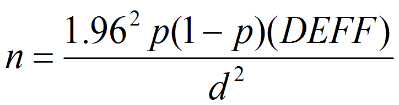

When	d	= Desired level of absolute precision = 1

DEFF	= Estimated design effect = 0.05

Thus	n = 189 + 38 (20% extra) = 227 patients/pregnant women

A decision was made to recruit 230 patient participants

Plus	60 medical personnel who volunteer to participate

**A total of 290 participants are required.**

14. **Study protocol**

Equipment

1. The Baiya’s Rapid Covid-19 IgM/IgG antibodies test kit
2. Applied Biosystems 7500 Real-time PCR system (Life Technologies, US) & the Novel Coronavirus (2019-nCoV) Nucleic Acid Diagnostic Kit (PCR-Fluorescence Probing) (SANSURE BIOTECH INC, Republic of China)

Protocol

1. Written informed consent is obtained from the participant
2. Participant information; including demographic data, diagnosis and type of operation (patients), risks of COVID-19 infection (i.e., history of contact, travelling to high risk area) , signs and symptoms of upper respiratory tract infection, was obtained
3. Blood sampling is performed by either venipuncture (for IV access) or fingertip collection for the Baiya’s Rapid COVID-19 IgM/IgG test kit.

*The test was performed according to the instruction manual. Briefly, 1-2 drops of blood are required and dropped onto the kit followed by 2-3 drops of dilution buffer. Result is read within 15 minutes.

4. Nasopharyngeal swab is performed and submitted for the real-time RT-PCR.
5. The RT- PCR for SARS-CoV-2 was performed on the ABI7500 real-time PCR machine (ThermoFisher, Waltham, USA) using a commercial kit (Sansure Biotech Inc, China).

In the emergency condition, the patient participant will undergo an operative procedure first, while the nasopharyngeal swab is obtained later (in PPE).

Medical personnel who take care of either ‘patient under investigation’, or participant who is at risk (history of contact with a confirmed case or having URI symptoms) for COVID-19 infection, or participant with the rapid test, will receive appropriate PPE and work in the negative environment. For other patients, protective equipment is depended on the clinician’s judgement.

15. **Outcome measurement / Data Analysis)**

**Primary outcomes**

Diagnostic values: Test sensitivity, specificity, false positive/negative rates, positive and negative predictive value, accuracy of the Rapid antibody test kit

**Secondary outcomes**

Sero-pravalence and Incidence of COVID-19 infection in the study population

ROC curves – Area under the curve (AUC) of both the rapid test and ‘patient under investigation criteria’

16. **Study Period**

Total study period: 6 months from July 2020 – Jan 2021

Data collection period: 2-3 month depending on the pandemic

17. **Reference**

1. Bai Y, Yao L, Wei T, et al. Presumed Asymptomatic Carrier Transmission of COVID-19. JAMA. Published online February 21, 2020. doi:10.1001/jama.2020.2565
2. (2020). FACT SHEET FOR HEALTHCARE PROVIDERS New York SARS-CoV-2 Real-time RT-PCR Diagnostic Panel.Fda.gov. Retrieved 13 March 2020, from <https://www.fda.gov/media/135662/download>.
3. Li Z, Yi Y, Luo X, Xiong N, Liu Y, Li S, et al. Development and Clinical Application of A Rapid IgM-IgG Combined Antibody Test for SARS-CoV-2 Infection Diagnosis. Journal of medical virology. 2020.
4. Sun P, Lu X, Xu C, Sun W, Pan B. Understanding of COVID-19 based on current evidence [published online ahead of print, 2020 Feb 25]. J Med Virol. 2020;10.1002/jmv.25722. doi:10.1002/jmv.25722
5. Breslin N, Baptiste C, Miller R et al. COVID-19 in pregnancy: early lessons,American Journal of Obstetrics & Gynecology MFM (2020) doi: https://doi.org/10.1016/j.ajogmf.2020.100111. 6
6. Liu D, Li L, Wu X et al. Pregnancy and perinatal outcomes of women with

coronavirus disease (COVID-19) Pneumonia: A preliminary analysis. AJR 2020:

215: 1-6. doi.org/10.2214/AJR.20.23072

7. Chen H, Gui J, Wang C et al. Clinical characteristics and intrauterine vertical

transmission of potential COVID-19 infection in nine pregnant women; a

retrospective review of medical records. Lancet 2020; 395: 809-815

8. คณะอนุกรรมการมาตรฐานวิชาชีพ. RTCOG Clinical Practice Guideline: Management of COVID-19 Infection in Pregnancy. Royal Thai College of Obstetricians and Gynecologist 2020, published 20 March 2020
9. Sutton D, Fuchs K, D’Alton M, and Goffman D. Universal Screening for SARS-CoV-2 in Women Admitted for Delivery: Letter to Editor (2020) Nejm published online April 2020, DOI: 10.1056/NEJMc2009316
